# Evaluating a first fully automated interview grounded in Multiple Mini Interview (MMI) methodology: results from a feasibility study

**DOI:** 10.1101/2021.02.28.21251817

**Authors:** Alison Callwood, Lee Gillam, Angelos Christidis, Jia Doulton, Jenny Harris, Marianne Coleman, Angela Kubacki, Paul Tiffin, Karen Roberts, Drew Tarmey, Doris Dalton, Virginia Valentin

## Abstract

**Objectives:** Global, Covid-driven restrictions around face-to-face interviews for healthcare student selection have forced admissions staff to rapidly adopt adapted online systems before supporting evidence is available. We have developed, what we believe is, the first fully automated interview grounded in Multiple Mini-Interview (MMI) methodology. This study aimed to explore test re-test reliability, acceptability and usability of the system.

**Design, setting and participants:** mixed-methods feasibility study in Physician Associate (PA) programmes from two UK and one US university during 2019 - 2020.

**Primary, secondary outcomes:** Feasibility measures (test retest reliability acceptability and usability) were assessed using intra-class correlation (ICC), descriptive statistics, thematic and content analysis.

**Methods:** Volunteers took (T1), then repeated (T2), the automated MMI, with a seven-day interval (+/− 2) then completed an evaluation questionnaire. Admissions staff participated in focus group discussions.

**Results:** Sixty-two students and seven admission staff participated; 34 students and four staff from UK and 28 students and three staff from US universities.

Good-excellent test-retest reliability was observed with T1 and T2 ICC between 0.62-0.81 (p<0.001) when assessed by individual total scores (range 80.6-119), station total scores 0.6-0.91, p<0.005, individual site (all ICC≥ 0.76 p<0.001) and mean test retest across sites 0.82 p<0.001 (95% CI 0.7-0.9).

Admissions staff reported potential to reduce resource costs and bias through a more objective screening tool for pre-selection or to replace some MMI stations in a ‘hybrid model’. Maintaining human interaction through ‘touch points’ was considered essential.

Users positively evaluated the system, stating it was intuitive with an accessible interface. Concepts chosen for dynamic probing needed to be appropriately tailored.

**Conclusion:** These preliminary findings suggest that the system is reliable, generating consistent scores for candidates and is acceptable to end-users provided human touchpoints are maintained. Thus, there is evidence for the potential of such an automated system to augment healthcare student selection processes.

## Introduction

Global, Covid-driven social distancing restrictions have forced healthcare admissions staff to rapidly adapt to online systems^1^. The rate of change has outstripped published evidence, resulting in interview methods with largely unknown efficacy. Our responsibilities to ensure inclusive and robust processes have therefore never been more challenging to enact. Pre-pandemic, candidate selection was predominantly face-to-face using unstructured or structures approaches including panel interviews, group interviews and multiple mini interviews (MMIs)^2^. MMIs are series of short, focused interactions with a number of different interviewers^3^. This multi-station format featuring scenario questions, tailored scoring pro forma and a unidirectional flow of conversation is designed to mitigate against the potential impact of interviewer bias^3^. MMI scenarios focus on random subject areas intended to assess role-defined attributes and values. This makes it more difficult for candidates to anticipate questions and benefit from any prior ‘coaching’ by preparing answers. None-the-less, MMIs along with other face-to-face methods are understood to be costly, resource intensive and influenced by unintended bias intrinsic to human assessment^2^.

Technology-facilitated interviews aim to alleviate such cost and bias issues. However, the limited evidence that is available evaluating examples in healthcare student selection, including Skype multiple mini interviews (MMIs) ^4^, asynchronous MMIs^5^ and asynchronous panel interviews^6^, is inconclusive. For example, candidates report feeling that the ability to fully express themselves is impaired while others consider the absence of an interviewer makes the process more objective^5^. Recently the challenges of conducting online interviews have been discussed^7,8^ but detailed evaluation data remains insubstantial.

Beyond health professions, multinationals describe resource and bias reduction achieved through technology-enhanced interviews. Unilever, for example, use artificial intelligence (AI) to analyse candidates’ interviews based on facial expressions and word choice. They report a 50% cost-reduction and 16% increase in diversity hiring^9^ due to improved accessibility to interviews and reduced opportunity for unconscious bias. There is insufficient evidence to draw causal inferences from these insights, but they remain potentially relevant to health professions selection where facilitating fairness is an international priority area^10^.

Nonetheless, pre-pandemic, the use of online technology in healthcare admissions was exploratory with efficacy yet to be formally established in a significant number of cohorts.

This paper presents preliminary evaluation of, what we believe is the first known automated interview grounded in MMI methodology, intended to improve cost-efficiency and reduce unintended bias associated with human assessments as well as overcome social distancing restrictions.

Our automated interview emulates the principles of face-to-face MMIs^3^ where interview content is analysed for the demonstration of role-defined values and attributes, but not by a human. An advanced, custom-built digital system combining validated ‘off-the-shelf’ and bespoke technologies uses techniques of natural language processing (text mining) to identify evidence of construct relevant attributes and values from narrative interview content. A minimum word limit is required to enable this in-depth analysis. Results provided to assessors are intended to help inform selection decisions where the ability to sense-check the reasons for allocated scores makes for a transparent decision-support tool. The system is summarised in Figure 1. The difference between our automated interview and currently adapted online interviews is that it provides for a fully automated interview as opposed to a person using videoconference technology to facilitate a human-assessed interview using MMI scenarios.

**Figure 1.**
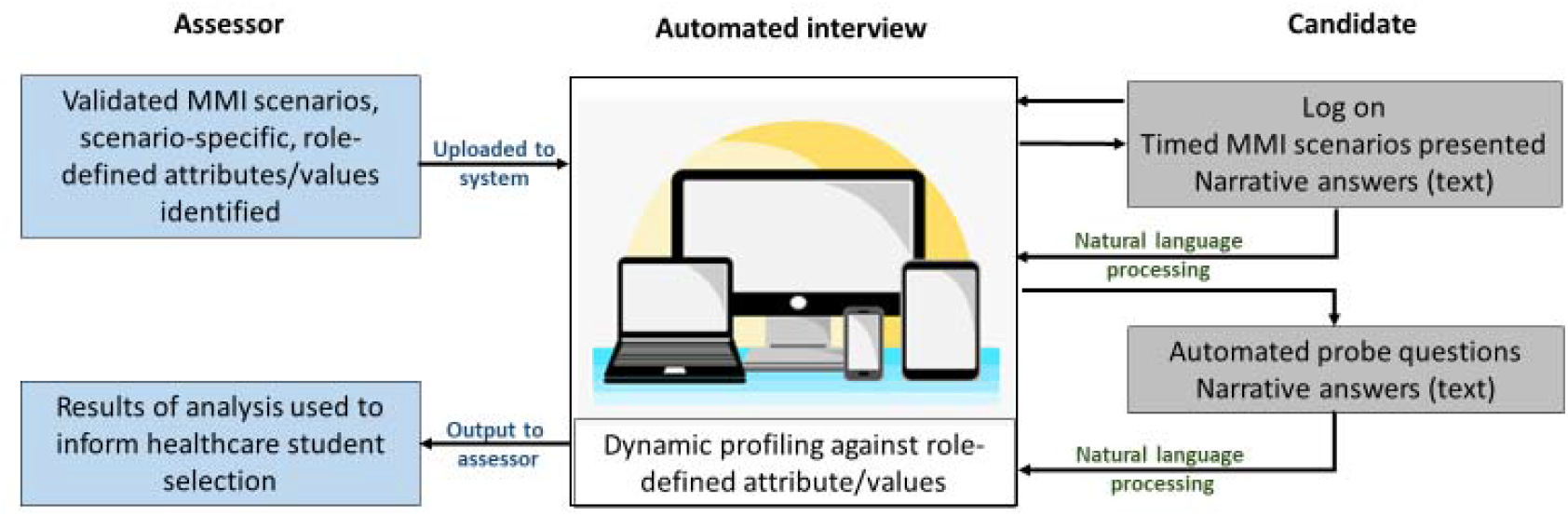
Diagrammatic representation of the automated interview.

## Methods

### Design

The development and evaluation of the automated interview, grounded in MMI methodology, took place in three phases; scoping, pre-test and feasibility study between January 2018-January 2020. This paper focuses on the outcomes of the feasibility study (April 2019 – December 2019) which aimed to evaluate the usability, acceptability and test re-test reliability of our automated interview system in admissions to Physician Associate (PA) programmes in two UK and one US university. For completeness, prior work is summarised in Table 1.

**Table 1.**
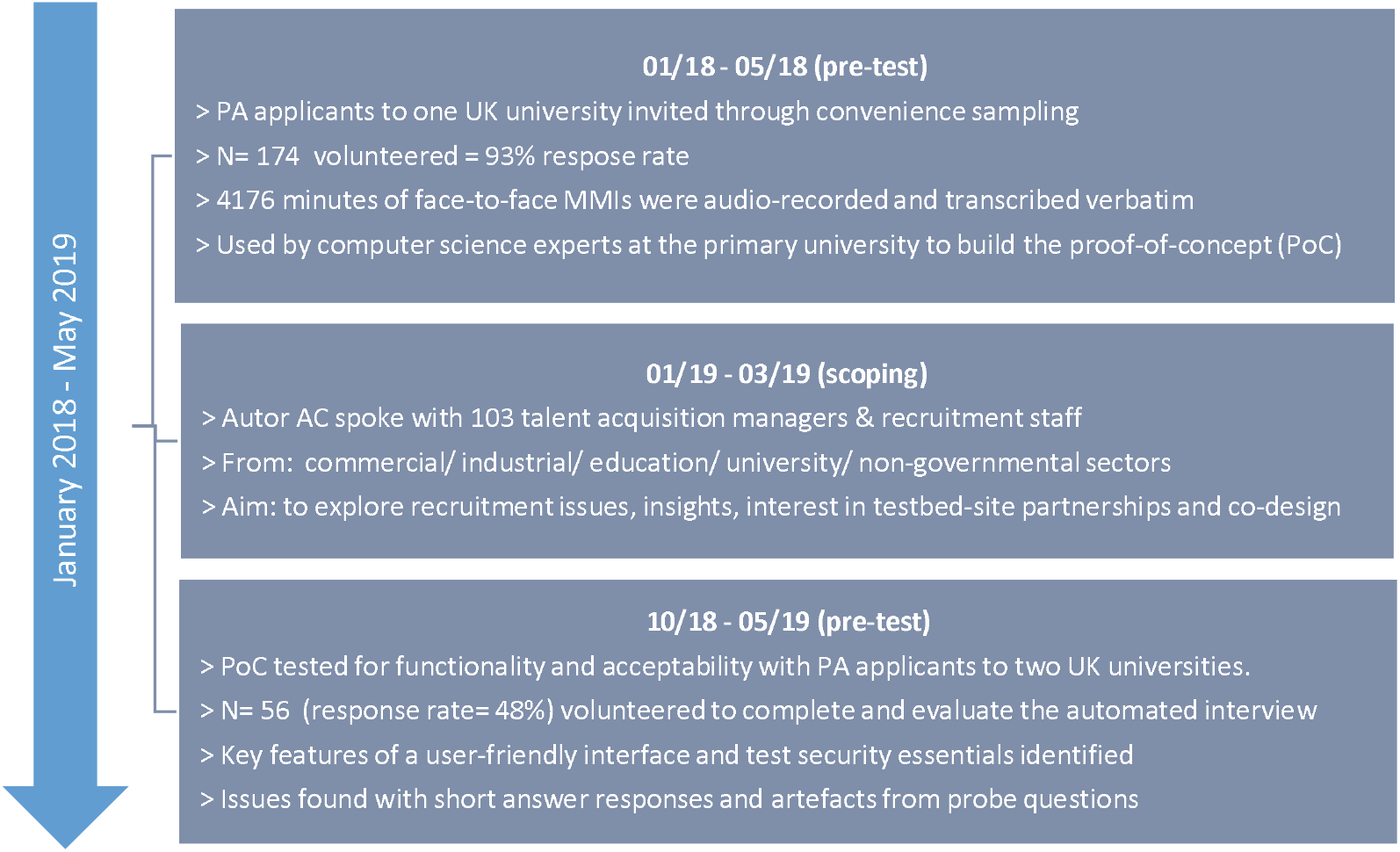
Scoping and pre-test activity.

This dual paradigmatic, dialectical enquiry^11^ was underpinned by Olsen and Eoyang’ Complex Adaptive Systems (CAS) model^12^. The pragmatic, ‘evolving’ systems approach enabled refinements to be made to the system from theoretical conception to deployable system where we were open to new insights as they emerged. The iterative nature of this model aligned with meeting the challenges of developing and piloting a new approach for which there was no known precedent.

### Participants

UK universities were invited to act as testbed sites using an MMI Expert Group network. In the US, an invitation was sent to PA programme admissions leads through a national network.

Admissions staff leading PA student selection at collaborating testbed sites worked with the research team to facilitate setting up the automated interview including supplying site-specific scenarios.

Volunteers to take the automated interview were recruited through non-probability convenience sampling from PA students at collaborating universities between April 2019 – December 2019. In the pre-pilot, applicants to PA programmes were invited to participate, but this brought challenges to applicants and staff on already stressful interview days. Therefore, study recruitment was broadened to include first year PA students. This approach aligned with the study aims because, at this stage, we were interested in test re-test performance against successive automated interview scores, deemed an essential step prior to validity testing with ‘live’ applicants.

Universities who already used MMIs and first year PA students at collaborating test-bed sites met inclusion criteria. Universities who did not use MMIs and PA students who had been involved in the scoping/pre-pilot development of the automated interview were excluded from the study.

### Data collection

PA students took part at a designated date, time, and venue with secure computer access and stable Wi-Fi. They completed four MMI-style scenarios using the automated interview system, writing their answers in text form, allowing a maximum 40 minutes overall, +/-10 minutes for additional time if required (T1). Using text responses (as opposed to oral) was a pragmatic decision taken at this stage as we were interested in the ‘real time’ capability of the system and wanted to be confident in capturing responses. Scenarios were site-specific, the content being replications of the face-to-face MMI scenarios used to interview students during their ‘live’ selection. For test re-test evaluation, volunteers were asked to repeat the same four scenarios one week later (± 2 days, T2) under similar conditions as T1, thereby minimising carry-over effect and the impact of any ‘learning’^13^.

Admissions staff participated in site-specific focus groups to elicit acceptability perspectives, defined according to Nielsen (1993) ^14^. An *a priori* topic guide facilitated exploration of their views of the system itself and automation in candidate selection.

To explore usability^14^ students completed a study-specific evaluation questionnaire immediately following T2. The questionnaire contained a mixture of closed questions with Likert scales and open text formats.

### Analysis

Automated interview scores for each of the attributes/values were summed at T1 and T2 for each candidate per station and across stations. Descriptive statistics were explored, and test retest reliability was assessed using the intraclass correlation coefficient (ICC) two-way mixed-model^15^. Individual total scores, station total scores, per site and mean scores for T1 and T2 are presented. All analysis was performed in Stata v16. Scores for attribute/value comparisons at T1 and T2 were also verified using multidimensional distance measures including Manhattan, Euclidean and Cosine measures^16^.

Staff focus group discussions were transcribed verbatim and thematic analysis^17^ performed by author XX, who was not otherwise involved in the study. This involved reading the transcripts in detail and multiple coding passes using NVivo 12 (QSR International, USA). Emerging themes were reviewed, and coding conflicts resolved collaboratively with research team member, author XX.

Descriptive statistics of students’ characteristics and views are presented. Qualitative content analysis^18^ was performed on open-ended questions to elicit students’ perspectives of the automated interview software usability.

## Results

### Sample characteristics

A total of 62 first year PA students from one US and two UK universities took part (UK1: n = 17; UK2: n = 17, US n = 28), representing 52% average uptake across sites.

English was the first language of over 70% of student participants. US students differed demographically from those at the two UK sites, with a more even distribution of age groups, lower proportion of females, and being predominantly white. In the UK universities, over 80% of participants were under 30 years of age, and over 59% were female with greater ethnic diversity. Volunteers had some prior exposure to pre-selection online assessment systems, including University Clinical Aptitude Test^19^ in the UK and CASPer^20^ in the US, but not a fully automated interview (Table 2).

**Table 2.** User Characteristics (n = 62 students)

|  |  | **US (n = 28)** | **UK1 (n = 17)** | **UK2 (n = 17)** |
| --- | --- | --- | --- | --- |
| **English as a first language** |  | 22 (78.6%) | 12 (70.6%)<br>Missing n = 1 | 14 (82.4%)<br>Missing n = 2 |
| **Gender** | Female | 12 (42.9%) | 10 (58.8%) | 15 (88.2%) |
| **Age Group** | Under 30 | 14 (50%) | 14 (82.4%) | 15 (88.2%) |
|  | 30 and above | 14 (50%) | 3 (17.6%) | 2 (11.8%) |
| **Ethnicity** | White | 15 (53.6%) | 7 (41.2%) | 5 (29.4%) |
|  | Asian | 2 (7.14%) | 7 (41.2%) | 3 (17.6%) |
|  | Black | 0 (0%) | 3 (17.6%) | 6 (35.3%) |
|  | Mixed/Other | 10 (35.7%) | 0 (0%) | 2* (11.8%) |
| **How helpful did you find the instructions? Median (IQR, skewness)** |  | 3.5 (1, -0.651) | 3 (1.5, -0.237) | 3 (0, 0.051) |
| **How intuitive was the system? Median (IQR, skewness)** |  | 4 (1.00, -0.796) | 3 (1.00, -0.115) | 3 (1.00, -0.855) |
| **How did probe questions relate to overall scenario presented at the beginning? Median (IQR, skewness)** |  | 3 (1, 0.584) | 2 (1, 1.035) | 2 (1.5, 0.054) |
| **Were the probes helpful in allowing you to expand on the answer? Median (IQR, skewness)** |  | 2 (0.75, 0.578) | 2 (1, 0.741) | 2 (0.5, 0.057) |
| **Timer** | Less than 3 minutes | 3 (10.7%) | 1 (5.9%) | 0 (0%) |
|  | 3 to 5 minutes | 13 (46.4%) | 9 (52.9%) | 3 (17.6%) |
|  | 5 minutes or more | 12 (42.9%) | 7 (41.2%) | 14 (82.4%) |
| **Previous experience of online assessment** |  | 13 (46.4%) | 2 (11.8%) | 3 (17.6%) |

Likert scales rated 1-4 with 1 representing a negative statement e.g. not helpful at all and 2 to 4 ranging from least positive e.g. sometimes helpful to most positive e.g. always helpful) \* Includes Other – Prefer not to say.

English was the first language of the seven participating admission staff. Five described themselves as White, one British Asian and one American Asian. There was a gender imbalance with six female participants and one male and all were over 40 years of age.

### Test retest evaluation

Complete data including automated interview scores were available for 57/62 (92%) participants (US n= 26, UK1 n = 14, UK2 n = 17). Two volunteers were unable to finish the retest in US for personal reasons; attrition at the other sites was due to incomplete/missing data.

Good-excellent reliability was demonstrated with T1 and T2 intra-class correlation (ICC) between 0.62-0.81, p<0.001 when assessed by individual total scores (range 80.6-119), station total scores between 0.6-0.91, p<0.005, individual site (all ICC≥ 0.76 p<0.001) and mean test retest across sites 0.82 p<0.001 (95% CI 0.7-0.9). Table 3. Manhattan, Euclidean and Cosine measures showed intra-candidate consistency was generally stronger than T1/T2 inter-candidate comparisons.

**Table 3.** Intraclass correlations between Test 1 and Test 2 per station, individual and across sites.

| **ICC per station (total scores) at T1 and T2 per site** |  |  |  |
| --- | --- | --- | --- |
| **Test bed site** | **ICC T1 and T2** | **95% CI** | **P value** |
| **US n=26** |  |  |  |
| Station 1 | .77 | .38-.87 | .001 |
| Station 2 | .60 | .06-.81 | .008 |
| Station 3 | .78 | .49-.89 | .000 |
| Station 4 | .75 | .45-.89 | .000 |
| **UK2 n=17** |  |  |  |
| Station 1 | .80 | .30-.87 | .001 |
| Station 2 | .79 | .44-.93 | .001 |
| Station 3 | .91 | .74-.97 | .000 |
| Station 4 | .74 | .29-.91 | .005 |
| **UK1 n=14** |  |  |  |
| Station 1 | .43 | -.79-.82 | .164 |
| Station 2 | .52 | -.49-.85 | .098 |
| Station 3 | .73 | .16-.91 | .012 |
| Station 4 | .02 | -.16-.73 | .374 |

| Test site | T1 mean (SD) | T2 mean (SD) | ICC of total scores per student (95% CI) | ICC average (95% CI) | P value |
| --- | --- | --- | --- | --- | --- |
| US n= 26 | 104.3 (5.49) | 102.9 (7.93) | 0.65 (0.35, 0.82) | 0.79 (0.52, 0.90) | <0.001 |
| UK2 n= 17 | 100.2 (7.84) | 99.7 (6.32) | 0.81 (0.55, 0.93) | 0.89 (0.71, 0.96) | <0.001 |
| UK1 n= 14 | 97.3 (6.91) | 99.2 (9.40) | 0.62 (0.15, 0.86) | 0.76 (0.27, 0.92) | 0.007 |
| All sites n= 57 | 101.4 (7.1) | 101.0 (7.9) | 0.70 (0.54, 0.81) | 0.82 (0.70, 0.90) | <0.001 |

### Acceptability

Seven admissions staff participated in three focus groups (US n = 3, UK1 n =2, UK2 n = 2) representing all those who were approached. The following key themes emerged from analysis of the discussions, illustrated in Table 4.

**Table 4.**
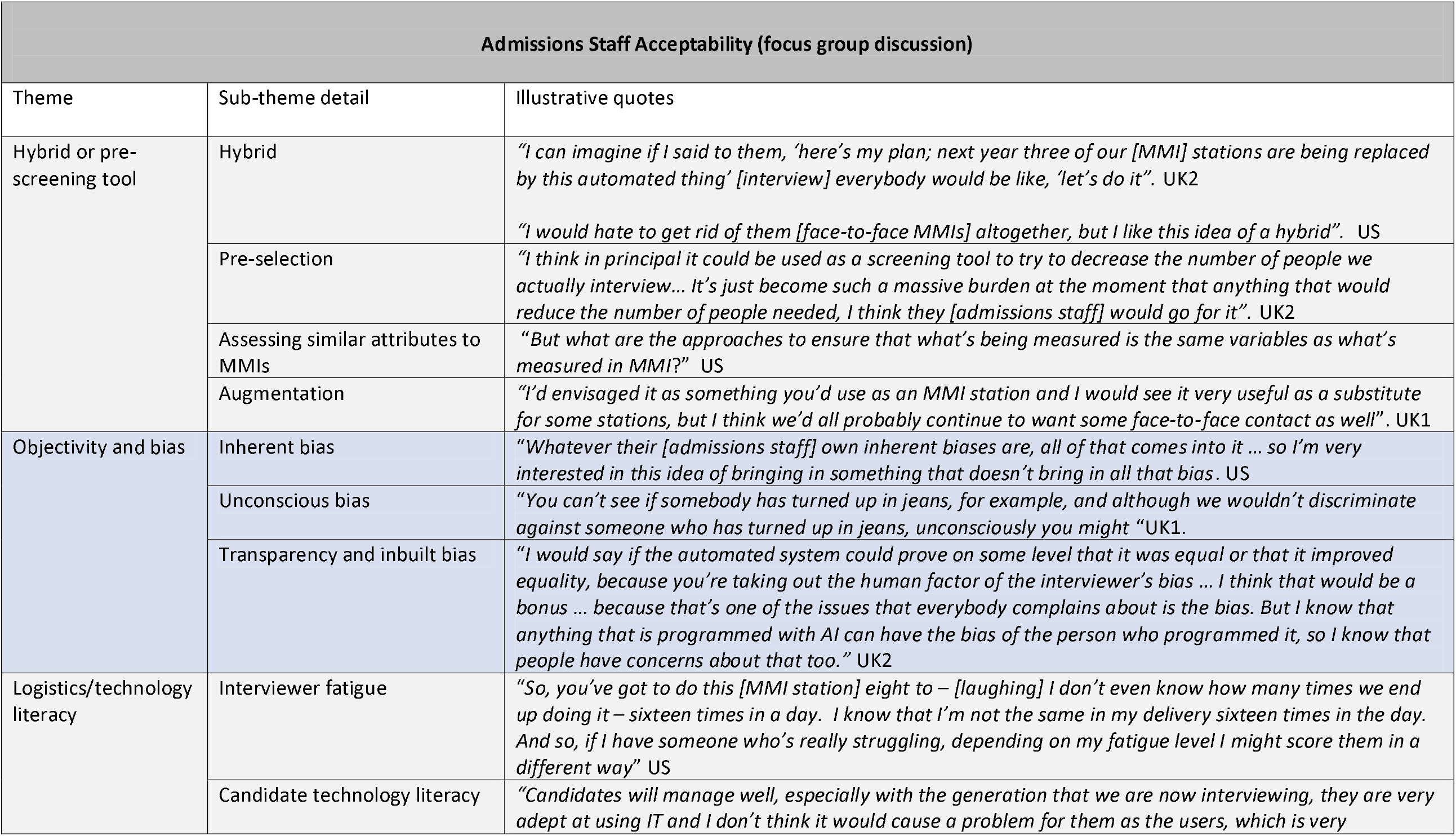
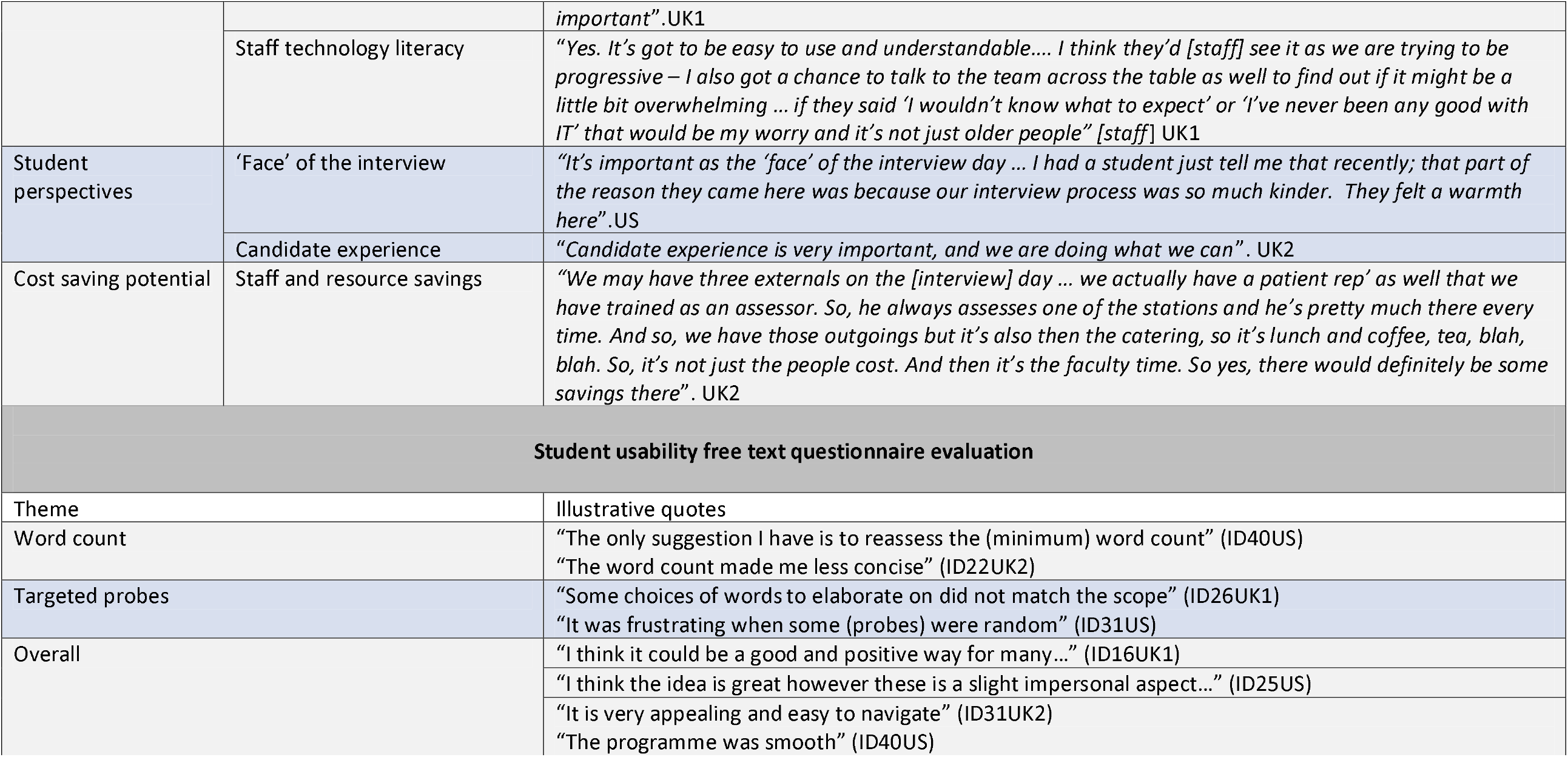
Acceptability and usability evaluation; key themes and illustrative quotes.

#### Hybrid or Screening tool

Admissions staff from all three universities felt the system could be adopted as an augmentation to in-person interviews in a hybrid approach. There was agreement that selection processes needed some degree of human involvement. It was suggested this could take several forms including: face-to-face contact with an interviewer for MMI stations; people to supervise and support the automated interview ensuring technical issues did not disadvantage anxious students; or as an opportunity for prospective students and faculty staff to meet one-another in person.

Admissions staff also saw a place for the automated interview as a pre-selection interview tool with potential to mitigate some of the resource costs of conducting face-to-face interviews like MMIs. It was thought that greater focus could then be directed at differentiating between those selected for final interview, reducing the number of interviews having to be conducted by staff.

All admissions staff felt it was important for the automated interview system to be able to measure the same or broadly similar candidate qualities as MMIs in order to be suitable as a direct replacement. This was due to an implicit trust in the ability of the face-to-face MMI methodology to enable optimal selection of the desired candidates.

#### Objectivity and Bias

Reducing subjectivity and bias were perceived by admissions staff across sites to be the core benefit and appeal of the system through consistency of evaluation and scoring. As a pre-screening tool, it was thought that interviewer burden from volume of MMIs, understood to exacerbate tiredness and increase the likelihood of bias emerging^21^, could be reduced.

#### Logistics

Admission staff felt applicants would manage well with the automated interview interface because of the increasingly widespread use of online selection processes for e.g. part-time jobs. Technology literacy concerns relating to the ability of staff to respond to queries arising with a new system were raised, particularly in the US site.

#### Student Perspectives

Admission staff across universities acknowledged that interviews can be stressful experiences and avoiding technical hiccups was an important priority. A positive applicant experience was thought to be essential. Some concerns were raised, particularly by UK1, that a computer-based interview may not be well received by applicants compared to a face-to-face interview thereby impacting their attitude towards the university.

### Usability

All students who participated in the study (n=62) went on to complete the post-automated interview evaluation questionnaire.

Students were positive overall (median score ≥3 across sites) about the user interface, instructions and the intuitiveness of the system stating it: ‘was very appealing’, ‘easy to use’ and ‘ran smoothly’ (Table 2).

When asked about the probe questions in terms of their relevance to the overall scenario and how helpful they were, responses were less positive with a median rating of 2 across all three sites, with the exception of the US university who were more positive. Open text questionnaire responses suggested that the concepts chosen for students to expand on in the dynamic probing were not always relevant to the scenario or their answer. Only 22.5% of volunteers (n = 14) across all sites reported they felt the probe questions were consistently tailored to their answers.

Volunteers felt that more rather than less time was needed to respond to each of the scenarios, with most indicating a minimum of three minutes was needed. Preferences varied by site with the majority of UK2 university students feeling they needed 5 minutes or longer.

47% - 59% of volunteers, across sites, wrote free text comments. Over half of these reiterated the need for targeted probe questions or suggestions for a reduced minimum word limit alongside positive feedback as illustrated in Table 4.

## Discussion

This is the first known automation and evaluation of an online interview grounded in MMI methodology. Our preliminary findings in UK and US settings provide evidence of good-excellent test retest reliability as well as acceptability and usability, as long as the system is deployed to augment and not replace human decision-making, and probe questions are appropriately targeted. These insights take on greater significance given the context of the current pandemic resulting in an enforced move to online systems in the absence of robust evidence.

Response rates were above those expected given data collection was pre-pandemic. This may be illustrative of an emerging acceptance of, or at least familiarity with, technology augmented interviews already prevalent in recruitment processes outside the field of healthcare.

Our online system provides for a fully automated interview as opposed to a human using videoconference technology to facilitate a human-assessed interview using MMI scenarios. Despite the concept of the automated interview being progressive, admissions staff saw substantial potential to mitigate subjectivity issues associated with human-led interviews through unintended bias and interviewer fatigue^22^. This was based on the consistency of the automated interview in contrast to the perceived nuanced differences between human interviewers conducting MMIs. Generating further evidence to support or refute this is needed. We do not underestimate the potential for inbuilt system biases and recognise essential adherence to best principles in the ethical deployment of trustworthy AI^23^. This is an extremely complex area and a theoretical discussion is outside the scope of this paper.

Admissions staff across test-bed sites were unequivocal that humans should make final decisions about candidate suitability. The automated interview system was considered an augmentation to face-to-face interviews designed for more consistent, less biased evaluation. We acknowledge concerns that a completely automated process with no human-led decision-making may bring unfairness and GDPR issues^24^. An automated interview is more remote and abstract from ‘real life’ and some candidates might find difficulty expressing themselves and communicating effectively without eye contact^25^. Conversely, online interviews have the potential to open up possibilities for applicants removing the need for travel costs and meeting dress code requirements, making selection more accessible and therefore fairer. These considerations become more significant in Covid-times where the pandemic is forcing adoption of online methods, sometimes without human touch points or conclusive evidence of fidelity, predictive validity or efficacy. A larger scale study is planned to evaluate potential differences in scoring between current adapted (online) MMIs and our fully automated interview to establish appropriate comparison methods, scoring approaches and predictive validity.

Admissions staff were very positive about the possibility of the automated interview to reduce resource costs. There is very limited economic evaluation of online automated interviews in the healthcare selection space that would support these views and further cost-effectiveness analysis would be beneficial. Outside the field of healthcare, multinationals espouse savings over 80% through online interviews, but we need to be cautious that selection decisions are defensible and do not end up as expensive litigation cases^26^.

How the automated interview was received by applicants mattered to admissions staff, highlighting the need for clear communication to manage expectations and foster optimal applicant performance. Admissions staff should consider how they can incorporate human ‘touch points’ in their online interviews especially as current social distancing restrictions mean personal face-to-face contact is not possible. These can be embedded into the candidate experience by facilitating opportunities to ask questions while online, either during or outside the interview, through virtual campus tours and live webinars/chats.

Student volunteers’ overall positivity about the usability of the automated interview is interesting in the context that in the UK, 88% were under the age of 30 and in the US, almost half had prior experience of online interviews. The iterative co-design, scoping and pre-piloting activity appears to have resulted in a system fit for purpose when deployed in an academic setting. The issue of irrelevant probe questions was concerning and reflects the complexity of an automated interaction. It has been addressed in subsequent iterations of our automated interview.

Suggestions to reduce the minimum word limit might impact on the reliability of the linguistic analysis. A new speech capture version of the automated interview now addresses this, as it appears candidates are more able to articulate their answers when spoken, thereby readily reaching the minimum word limit.

### Study limitations

The sample size is small, particularly from UK1, which may have impacted on the findings^27^ and is limited to three universities. Student volunteers were self-selected and over 70% had English as their first language. Assessing for potential differential attainment in the automated interview for those with English as a first language, compared to those for whom it is not, requires a larger sample size and will be explored in the planned large-scale testing of the automated interview. Notably, different sites used different scenarios, so caution is needed when interpreting the combined ICC. However, it is reassuring that ICCs by site were similarly high i.e. all >=0.7.

### Study rigour

Author XX conducted the focus groups given her prior experience. A structured interview proforma was used to facilitate discussions to minimise deviation and potential bias. Audio-recordings were transcribed verbatim and 20% double-checked by the research team where 98% accuracy was found. Coding conflicts were resolved with input from XX, and final themes and subthemes were by agreement with all authors.

## Conclusion

At the time of writing, lack of evidence means the efficacy of current improvised online interviews is largely unknown. These preliminary findings suggest that our automated interview system is reliable, generating consistent scores for candidates and is acceptable to end-users. There is evidence for the potential of such a system to augment candidate selection, though the perceived importance of maintaining human input was highlighted. These valuable insights are applicable across health professions selection. Further research will focus on evaluating the validity of the automated scores generated against construct-relevant outcomes.

Our system significantly advances technology augmented interviews from videoconference-facilitated to a fully automated interview designed to assist admissions staff in making decisions about accepting or rejecting applicants. Conceptually, using technology in this way maybe a step too far for some but a welcome innovation for others. Nonetheless, a symbiotic relationship between humans and technology has been forced by social distancing restrictions and we should be open to understanding possible benefits as well as risks when facing an unknown future.

## Data Availability

The datasets used and/or analysed during the current study are available from the corresponding author on reasonable request. Detailed technical information is withheld due to commercial sensitivity.

## Acknowledgements

Grateful thanks to the Physician Associate students and collaborating universities who took part.

## Declarations

### Author contributions

AC, LG and Ach contributed to the technology development, study design, data collection, analysis, drafting and revision of this paper. JD, DD, DT, KR, AK, VV, PT contributed to the data collection, drafting and revision of this paper. MC and JH contributed to the data analysis, drafting and revision of this paper. All authors reviewed the final manuscript.

### Ethics

All participants gave informed consent. Ethical approval was given from all participating sites including:

- The primary site, The University of Surrey Research Ethics Committee (UEC/2017/111/FHMS).
- St Georges University of London Joint Research and Enterprise Services (title used as reference, no number allocated).
- University of Utah Research Integrity and Compliance Committee IRB, reference: 00125158.

